# Coverage and factor associated to the uptake of Intermittent preventive treatment for malaria during pregnancy (IPTp) in Cameroun in 2018: an analysis from the 2018 Cameroon Demographic Health Survey data

**DOI:** 10.1101/2022.10.12.22281001

**Authors:** Dominique Guimsop Ken, Ange Faustine Kenmogne Talla, Haoua Kodji, Jerome Ateudjieu

**Affiliations:** Departement of public health, Faculty of Medicine and Pharmaceutical Sciences, University of Dschang, Dschang, Cameroon

## Abstract

Malaria in pregnancy is a major public health issue, contributing to significantly increasing mother and children mortality and morbidity. Intermittent preventive treatment of malaria using Sulfadoxine-Pyrimethamine (IPTp-SP) is one of the key interventions recommended by WHO and in Cameroon to reduce the morbidity of Malaria in Pregnancy. The present study aims to assess the distribution and determinants of poor uptake of IPTp-SP (< 3 doses) among pregnant women in Cameroon.

We conducted a secondary analysis of data extracted from the Cameroun Demographic Health Survey conducted in 2018. Data were collected using a questionnaire administered in face to face to mothers with at least one child under 5, selected using a 2-stage stratified sampling process. Frequencies of mothers exposed to less than 3 doses of IPTp-SP were estimated with a 95% CI. A case-control design was used to assess the association between key suspected determinants and being exposed to less than 3 doses of IPTp-SP. This was measured by estimating crude and adjusted Odd-Ratios (ORs).

A total of 13527 childbearing age women were interviewed, of which 5528 (40.9%) met our selection criteria. Among those, 845 (15.3%) had not attended any Antenatal care (ANC) visit, 1109 (20%) had attended 1 to 3 visits, 3379 (61.1%) had attended 4 to 7 visits and only 195 (3.5%) had attended at least 8 visits. In all, 3398 (61.5%, CI: 60.2-62.8) had received less than 3 doses of IPTp-SP. Maternal age below 26 years (aOR=1.17, CI: 1.01-1.35), residence in an area of lower malaria endemicity (aOR=1.26, CI: 1.00-1.58), residence in the Sahelian regions (aOR=5.81, CI: 2.46-13.69), and having attended less than 4 ANC visits (aOR=1.30, CI: 1.08-1.57) were predictors of poor uptake of IPTp-SP. Conversely, residence in major cities (aOR=0.65, CI: 0.50-0.84), having attended a first ANC visit at 3 months or less of pregnancy (aOR=0.66, CI: 0.57-0.78) and having been followed on ANC by a medical doctor (aOR=0.22, CI: 0.13-0.35), were preventing factors for poor uptake of IPTp3+. Highest level of education (aOR= 1.10, CI: 0.90-1.32) was not found associated with the uptake of IPTp-SP.

Therefore, only a third of pregnant women in Cameroon take an optimal dose of IPTp-SP. This proportion vary widely with the place of residence, being significantly low in the Sahelian regions. Interventions to address identified determinants of low coverage of IPTp-SP like maternal age below 26 years, ANC characteristics or being resident of area with lower malaria endemicity ought to be tested.

## Introduction

Malaria, caused in humans by five species of *Plasmodium*, remains one of the most widespread infectious diseases. The heavy burden is mainly related to *Plasmodium falciparum* and *Plasmodium vivax*. It threatens nearly half of the world’s population, resulting in 229 million cases and 409.000 deaths in 2019, Africa accounting for over 90% of cases [1]. Malaria is significantly known to increase the risk of maternal and fetal anemia, abortion, low birth weight, and neonatal death [2-5]. To reduce the burden of the disease, efforts are channeled into 3 main strategies: Intermittent Preventive Treatment in pregnancy with Sulfadoxine-Pyrimethamine (IPTp-SP), use of insecticide-treated nets, and effective case management in all areas of moderate to severe transmission [6]. IPTp-SP policy recommends that each pregnant woman receive at least three doses of Sulfadoxine-Pyrimethamine (SP) throughout her pregnancy, each at least one month apart, which can be safely administered from 13 weeks to delivery [7, 8].

Despite implemented interventions, the 3 doses IPTp-SP coverage in 2020 was 34% overall in the 33 countries that adopted the preventive measure [1]. The poor performance can be explained by limited access to healthcare, poor training of health care providers, stock-outs of the drugs, maternal and pregnancy-related factors as level of education and multigravidity among others [9-12].

In Cameroon, the Ministry of Health adopted a new policy of chemoprevention of malaria since May 2002 as part of ANC through the use of chloroquine, which was later replaced in January 2004 by IPTp-SP following related WHO recommendations [13]. Available literature estimated in 2018 the 3 doses coverage of IPTp-SP among pregnant women in Cameroon between 23.5% and 54.9% depending on socio-demographic characteristics, antenatal care performance, and study type [9, 14]. These figures lag clearly behind the 80% coverage rate targeted by the country for 2023. No study has been conducted to understand limitation of health care systems in achieving its objectives. While such finding would help policymakers in their effort to improve the effectiveness of actual strategies or propose new ones. The present study evaluated the distribution of poor uptake of ≥3 doses of IPTp-SP among pregnant women in Cameroon and assessed its determinants.

## Methodology

### Study design and setting

We conducted a secondary data analysis of the 2018 Cameroon Demographic and Health Survey (DHS) [14]. The data were collected from a cross-sectional household-based survey targeting 22 territorial units of Cameroon including 10 regions (each parted into their respective urban and rural areas) and the two major cities of the country. One of the areas affected by the ongoing socio-political crisis was excluded. Household were selected by stratified cluster random sampling process and in each visited household the data were collected using a face-to-face administered questionnaire.

### Study population

The target population was all women aged 15–49 years residing in the selected cluster and biological mother of a living child aged less than at the day the selected household is visited.

### Participants extraction from the data base

From the variable “woman of childbearing age’’, we deducted the variable “mother of a under 5 years old child” by coding. From those selected we excluded participants who did not have recorded data on the age of the child, those who did not have data on whether or not they attented ANC visits and those did not have data on whether or not they took sulphadoxine pyrimethamine during the pregnancy of the child (Fig 1).

**Fig 1.**
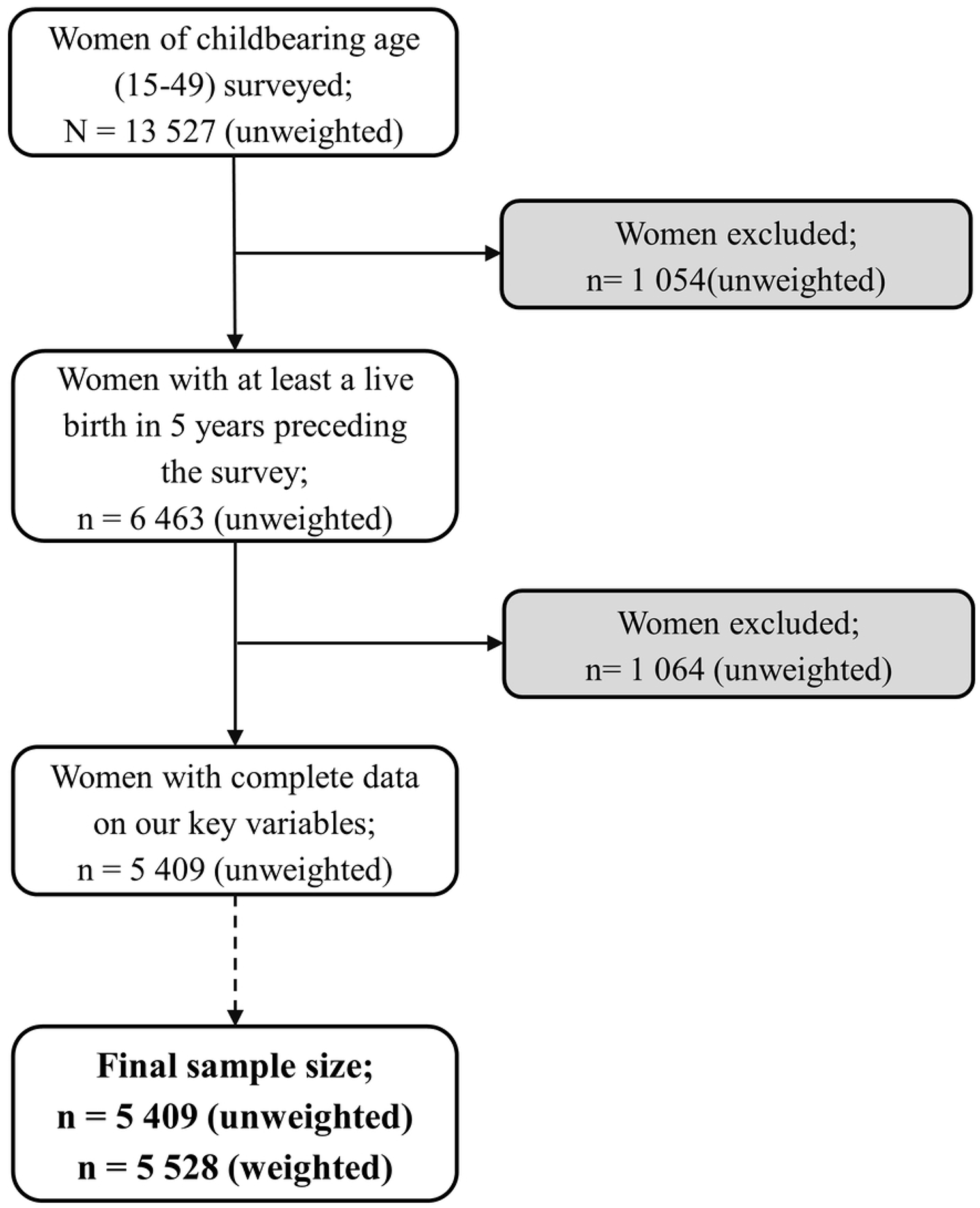
Flow diagram of sample selection from CDHS 2018 data.

### Variable of interest

Data were extracted for analysis based on variables that were theoretically and empirically linked to the uptake of ≥3 doses of IPTp-SP as follow: (a) the dependent variable was the number of women who had received less than three doses of IPTp-SP (Poor uptake of ≥3 doses of IPTp-SP) ; (b) Independent variables comprised woman’s age, stay in the place of residence, woman’s highest educational level, religion, literacy, wealth, health insurance coverage, occupational status, matrimonial status, partner’s highest level of education, region, area of residence, area’s malaria prevalence, wanted pregnancy, parity, history of a terminated pregnancy, ANC highest attendant, number of ANC visits. Variables regarding pregnancy and prenatal care were relative to the most recent live birth.

### Data management

Data extraction, grooming, and analysis were done using IBM SPSS Statistics version 26 (International business machines corp., New York, USA).

The complex sampling design of the survey was taken into account by using the “Complex samples” functionality of IBM SPSS for the analyses, and except specified otherwise, only weighted data were presented in our results.

### Statistical analysis

Categorical variables were summarized as frequencies and proportions. In analytic analysis, the Rao-Scott correction to Chi-square test was used to compare the distribution of the independent variables between the two levels of our dependent variable. Bivariate logistic regression was then used after all dependent variables were dichotomized, to measure the association with our independent variable. Dependents that had an association with the outcome significant at p<0.05 were selected for the multivariate logistic regression model. This estimated adjusted odds ratio (aOR) and their significance, controlling for the effect of other dependent variables. Collinearity was checked using Variance inflation factors (VIFs) calculation. Finally, substantive cross-level interactions among our dependents were explored, and only one was found significant, which was kept in the model. The level of significance used was 5% (0.05), two-tailed.

### Ethical consideration

The Cameroon DHS protocol, including measuring procedures and biological tests, was reviewed and approved by the Cameroonian ministry of health National ethical committee for research and human health (CNERSH) and Inner-City Fund (ICF) Institutional Review Board. Informed consent for the survey was obtained from each respondent before each interview. Permission to access the Cameroonian DHS 2018 survey dataset for the present study was obtained from the Demographic and Health Surveys (DHS) Program. Surveyors maintained confidentiality by an anonymization technique attributing a code to every case. The accessed data set was kept on a password-secured computer.

## Results

### Characteristics of the population

We included 5528 participants from the database, with almost half (48.1%, n=2657) aged between 25 and 35 years old and 44.5% (n=2455) having attended at least secondary school. About half (52.6%, n=2911) of the women lived in rural areas, and they seem to be fairly distributed across wealth classes. Overall, 61.5% (n = 3398) of the study population had taken less than three doses of IPTp-SP (Table 1).

**Table 1.** Socio-demographic and environmental characteristics of participants and comparison between those who took less than 3 doses of IPTp-SP and others (N=5528)

| Variables | Total<br>N=5528 | IPTp-SP uptake, n (%) |  | Rao-Scott F(df1, df2) | p |
| --- | --- | --- | --- | --- | --- |
|  |  | <3 doses<br>n=3398 | 3+ doses<br>n=2130 |  |  |
| Age groups (years) |  |  |  |  |  |
| 15-19 | 521 (9.4) | 333 (9.8) | 188 (8.8) | 0.93 (4.53, 1846.71) | 0.457 |
| 20-24 | 1205 (21.8) | 760 (22.4) | 445 (20.9) | . |  |
| 25-29 | 1496 (27.1) | 918 (27.0) | 578 (27.1) |  |  |
| 30-34 | 1161 (21.0) | 703 (20.7) | 459 (21.5) |  |  |
| 35-39 | 727 (13.1) | 421 (12.4) | 305 (14.3) |  |  |
| 40-49 | 416 (7.5) | 262 (7.7) | 155 (7.3) |  |  |
| Stay in the place of residence |  |  |  |  |  |
| Visitor | 121 (2.2) | 79 (2.2) | 42 (2.0) | 0.26 (2.83, 1158.50) | 0.844 |
| 0-19 years | 3454 (62.5) | 2119 (62.4) | 1334 (62.6) |  |  |
| 20-40] years | 219 (4.0) | 140 (4.1) | 79 (3.7) |  |  |
| Always | 1734 (31.4) | 1060 (31.2) | 674 (31.7) |  |  |
| Highest educational level |  |  |  |  |  |
| No formal education | 1425 (25.8) | 996 (29.3) | 429 (20.2) | 16.24 (2.77, 1130.29) | <0.001 |
| Primary | 1647 (29.8) | 1058 (31.1) | 590 (27.7) |  |  |
| Secondary | 2121 (38.4) | 1185 (34.9) | 936 (43.9) |  |  |
| Higher | 334 (6.1) | 160 (4.7) | 175 (8.2) |  |  |
| Religion |  |  |  |  |  |
| Animism/other | 212 (3.8) | 145 (4.3) | 67 (3.1) | 4.70 (1.97, 805.91) | 0.010 |
| Christian | 3786 (68.5) | 2246 (66.1) | 1540 (72.3) |  |  |
| Muslim | 1530 (27.7) | 1007 (29.6) | 523 (24.6) |  |  |
| Illiterate | 2016 (36.5) | 1391 (40.9) | 625 (29.3) | 30.88 (1.0, 408.0) | <0.001 |
| Combined wealth index |  |  |  |  |  |
| Poorest | 1111 (20.1) | 779 (22.9) | 331 (15.5) | 22.38 (3.75, 1531.02) | <0.001 |
| Poorer | 1215 (22) | 842 (24.8) | 374 (17.5) |  |  |
| Middle | 1135 (20.5) | 734 (21.6) | 401 (18.8) |  |  |
| Richer | 1100 (19.9) | 587 (17.2) | 514 (24.1) |  |  |
| Richest | 967 (17.5) | 457 (13.5) | 510 (23.9) |  |  |
| Health insurance coverage | 99 (1.8) | 45 (1.3) | 54 (2.5) | 7.62 (1.0, 408.0) | 0.006 |
| Current occupation | 3769 (68.3) | 2306 (67.9) | 1463 (68.7) | 0.21 (1.0, 408.0) | 0.650 |
| Ever in union | 4448 (80.5) | 2767 (81.4) | 1681 (78.9) | 3.74 (1.0, 408.0) | 0.054 |
| Partner's highest level of education, secondary + | 3636 (65.8) | 2336 (68.8) | 1299 (61) | 21.87 (1.0, 408.0) | <0.001 |
| Region |  |  |  |  |  |
| Sahelian regions | 2025 (36.6) | 1326 (39) | 699 (32.8) | 20.0 (1.94, 793.42) | <0.001 |
| Southern regions | 2528 (45.7) | 1611 (47.4) | 917 (43.0) |  |  |
| Douala/Yaounde | 975 (17.6) | 461 (13.6) | 515 (24.2) |  |  |
| Rural area of residence | 2911 (52.6) | 2000 (58.9) | 911 (42.8) | 48.75 (1, 41) | <0.001 |
| Area's malaria prevalence (%) |  |  |  |  |  |
| 0-9 | 1008 (18.2) | 560 (16.5) | 448 (21) | 508.0 (2.82, 1150.09) | 0.002 |
| 10-19 | 1717 (31.1) | 1061 (31.2) | 656 (30.8) |  |  |
| 20-29 | 1529 (27.7) | 907 (26.7) | 622 (29.2) |  |  |
| 30 + | 1274 (23.1) | 870 (25.6) | 404 (19) |  |  |

IPTp-SP, Intermittent preventive treatment in pregnancy with Sulphadoxine pyrimethamine

Up to 845 (15.3 %) women did not attend any ANC visit. Among the 3574 (64.6 %) women who attended the recommended number of visits, 1686 (47.2%) took ≥3 doses of IPTp-SP (Fig 2).

**Fig 2.**
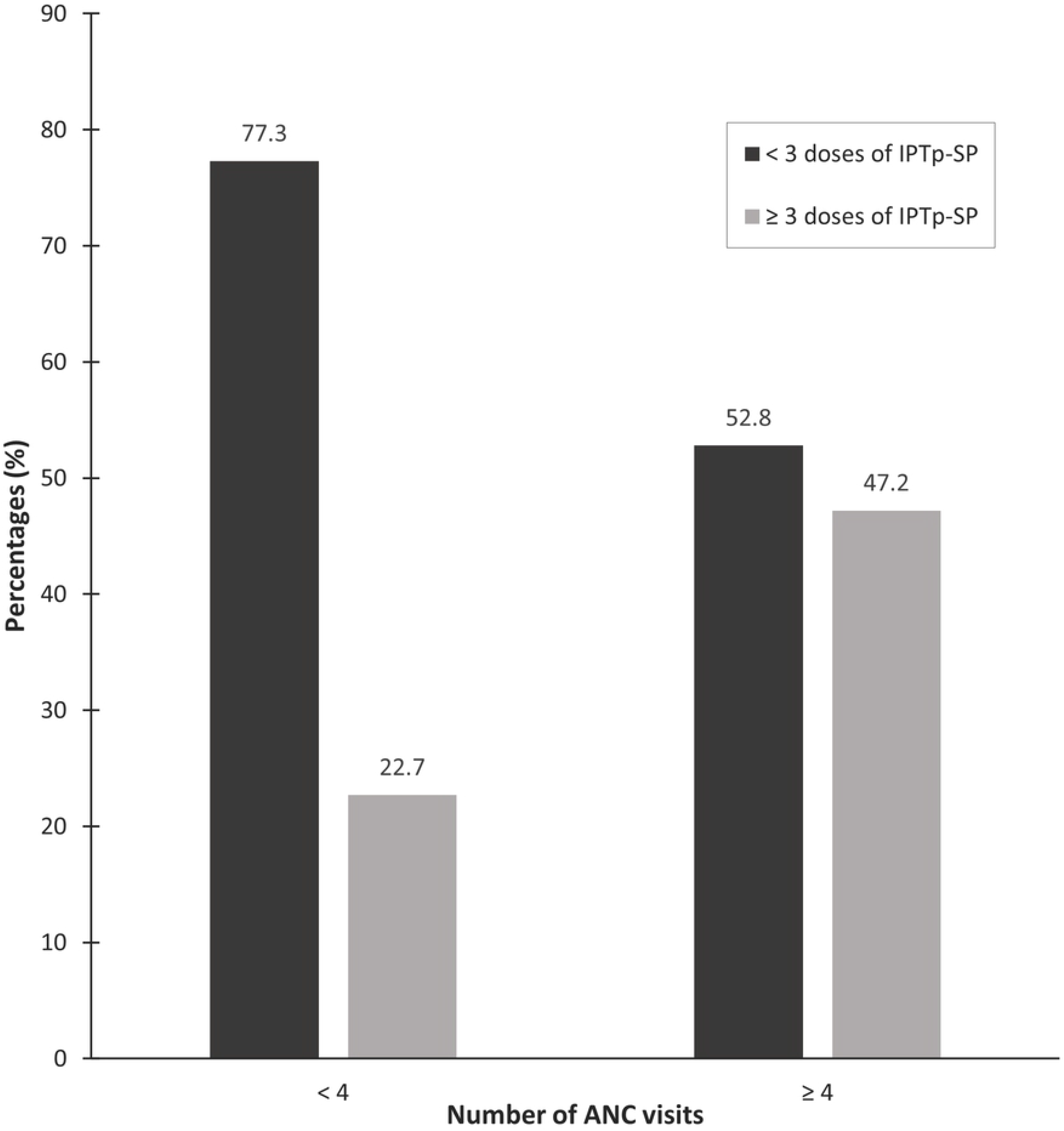
Proportion of uptake of ≥3 doses of IPTp-SP by number of ANC visits.

### Socio-demographic characteristics of participants and uptake of ≥3 doses of IPTp-SP

There was no association between the stay in the place of residence and uptake of IPTp-SP (p = 0.844). A lower proportion of women with poor uptake of IPTp-SP had attended secondary education compared to those who took more doses (39.6% vs. 52.1%, p ≤ 0.001). Furthermore, there were more illiterate among participants with poor uptake of IPTp-SP (40.9% vs. 29.3%, p < 0.001). Only 1.8% (n = 99) of pregnant women were covered by some health insurance; these were more represented among those who took ≥3 doses of IPTp-SP (p = 0.006). A lower proportion of women with poor uptake of IPTp-SP had ever been in union with a partner than women who received more doses (78.9% vs. 81.4%), though this association was not statistically significant (p=0.054). Moreover, more women among those with poor uptake of IPTp-SP had a partner with a secondary or more level of education (68.8% vs. 61%, p < 0.001). The Sahelian regions were more represented among those who took less than recommended 3 doses of IPTp-SP (39% vs. 32.8%) compare to those who took recommended doses. The distribution of women across areas with varying levels of endemicity was statistically different according to the uptake of IPTp-SP (p = 0.002).

### Obstetrical history of participants and uptake of ≥3 doses of IPTp-SP

Fewer women among those with poor uptake of IPTp-SP had a parity below 6 than among those who took more doses (p = 0.033). Similarly, fewer women with poor uptake of IPTp-SP had a history of terminated pregnancy (p = 0.022). There was a significant association between highest ANC attendant and uptake of IPTp-SP (p < 0.001), with a greater proportion of women among those with poor uptake of IPTp-SP having received no care (24% vs. 1.4%) and a lesser proportion having been cared by a doctor (22.2 vs. 34.9). Only 41.1 % of pregnant women had attended their first ANC visit before the fourth month of pregnancy; those were more represented among women who took recommended doses of IPTp-SP (54.5% vs. 32.8%, p < 0.001). The majority of women (61.1%) had attended 4 to 7 ANC visits. The association between the number of ANC attended and the uptake of IPTp-SP was also significant (p < 0.001) and a greater proportion of the women who took recommended doses had attended over 4 visits (79.2% vs. 55.5%). (Table 2)

**Table 2.** Obstetrical history and comparison between participants who took less than 3 doses of IPTp-SP and others (N=5528)

| Variables | Total<br>N=5528 | IPTp-SP uptake, n (%) |  | Rao-Scott F(df1, df2) | p |
| --- | --- | --- | --- | --- | --- |
|  |  | <3 doses<br>n=3398 | 3+ doses<br>n=2130 |  |  |
| Wanted pregnancy | 4250 (76.9) | 2585 (76.1) | 1665 (78.2) | 1.81 (1, 408) | 0.180 |
| Parity $\leq 5$ | 4432 (80.2) | 2682 (78.9) | 1750 (82.1) | 4.56 (1, 408) | 0.033 |
| History of terminated pregnancy | 1023 (18.5) | 584(17.2) | 439 (20.6) | 5.29 (1, 408) | 0.022 |
| ANC highest attendant |  |  |  |  |  |
| No care | 845 (15.3) | 816 (24) | 29 (1.4) | 93.30 (2.75, 1123.61) | <0.001 |
| Doctor | 1495 (27) | 753 (22.2) | 742 (34.9) |  |  |
| Nurse/midwife | 3012 (54.5) | 1734 (51) | 1278 (60) |  |  |
| Others | 176 (3.2) | 95 (2.8) | 80 (3.8) |  |  |
| 1<sup>st</sup> ANC visit before/at 3 months | 2274 (41.1) | 1114 (32.8) | 1160 (54.5) | 172.05 (1, 408) | <0.001 |
| Number of ANC visits |  |  |  |  |  |
| 0 | 845 (15.3) | 816 (24) | 29 (1.4) | 67.40 (4.61, 1880.81) | <0.001 |
| 1 | 83 (1.5) | 59 (1.7) | 24 (1.1) |  |  |
| 2 | 233 (4.2) | 167 (4.9) | 66 (3.1) |  |  |
| 3 | 793 (14.3) | 469 (13.8) | 324 (15.2) |  |  |
| 4 - 7 | 3379 (61.1) | 1792 (52.7) | 1586 (74.5) |  |  |
| 8 + | 195 (3.5) | 95 (2.8) | 100 (4.7) |  |  |

IPTp-SP, Intermittent preventive treatment in pregnancy with Sulphadoxine pyrimethamine; ANC, Antenatal care.

### Determinants of the uptake of less than three doses of IPTp-SP

Table 3 shows the results of logistic regression analyses. It indicates that independent predictors of poor uptake of IPTp-SP were age below 26 (p =0.038), residing in an area of lower endemicity (p = 0.047), in the Sahelian regions (p < 0.001) and having attended less than 4 ANC visits (p = 0.006). Women residing in large cities (p = 0.001), who attended a first ANC visit at 3 months or less of pregnancy (p < 0.001) and had been followed on ANC by a medical doctor (p < 0.001), were less likely to receive less than 3 doses of IPTp-SP than those residing in the southern regions, who attended first ANC visit latter and followed on by other health care workers. Last, the effect of the cross-level interaction between the ANC highest attendant and the Sahelian regions was positive and statistically significant (p < 0.001), indicating that the effect of the ANC highest attendant on the risk of a woman receiving less than recommended doses is conditional on whether the woman resides in the Sahelian regions; notably, women attended by medical doctors in the Sahelian regions had an even lesser risk of poor IPTp-SP uptake than those residing in the Southern regions (Fig 3).

**Fig 3.**
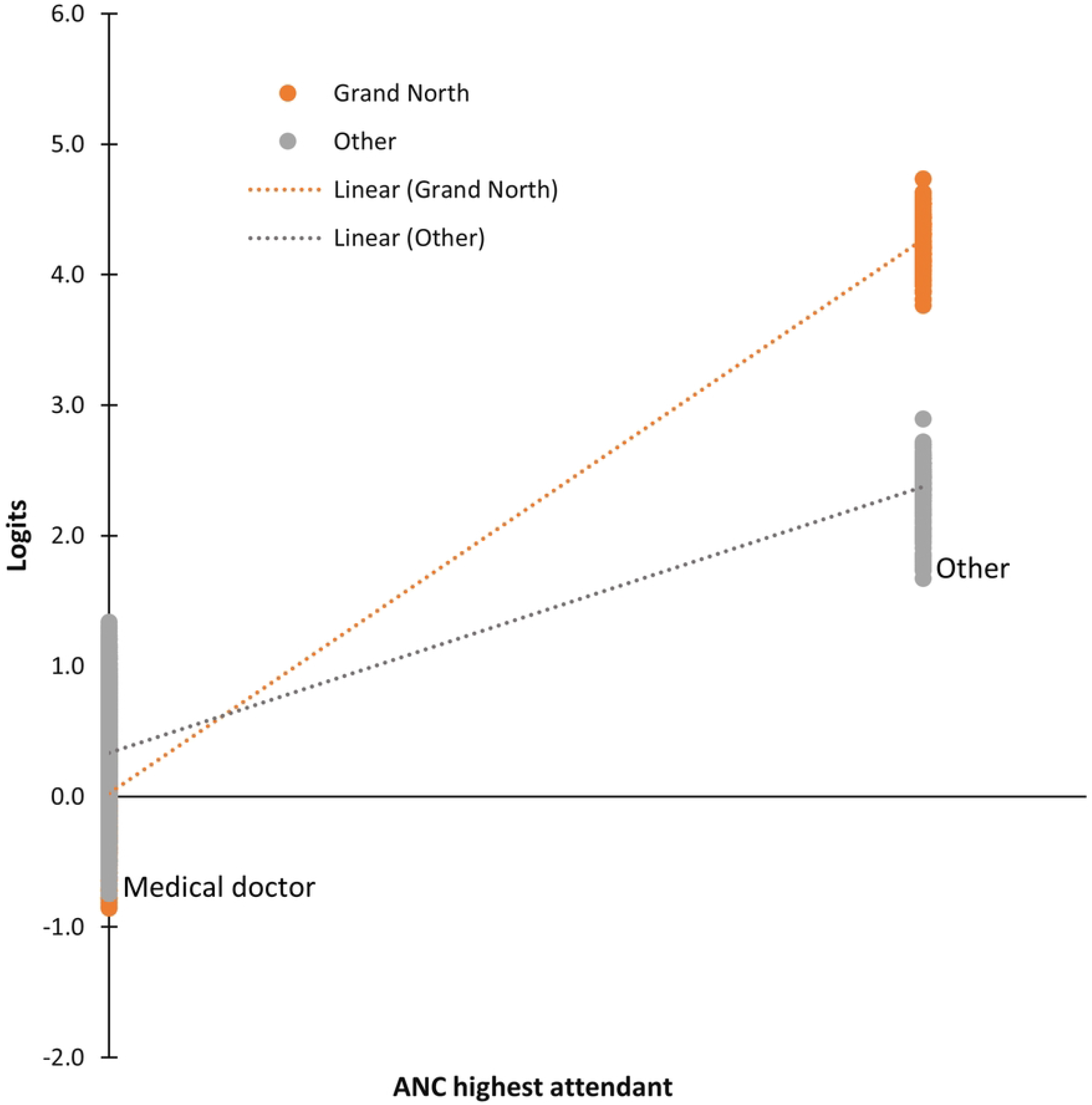
Cross-level interaction (ANC highest attendant x Sahelian regions region) plot.

#### Determinants of the uptake of less than 3 doses of IPTp-SP by pregnant women

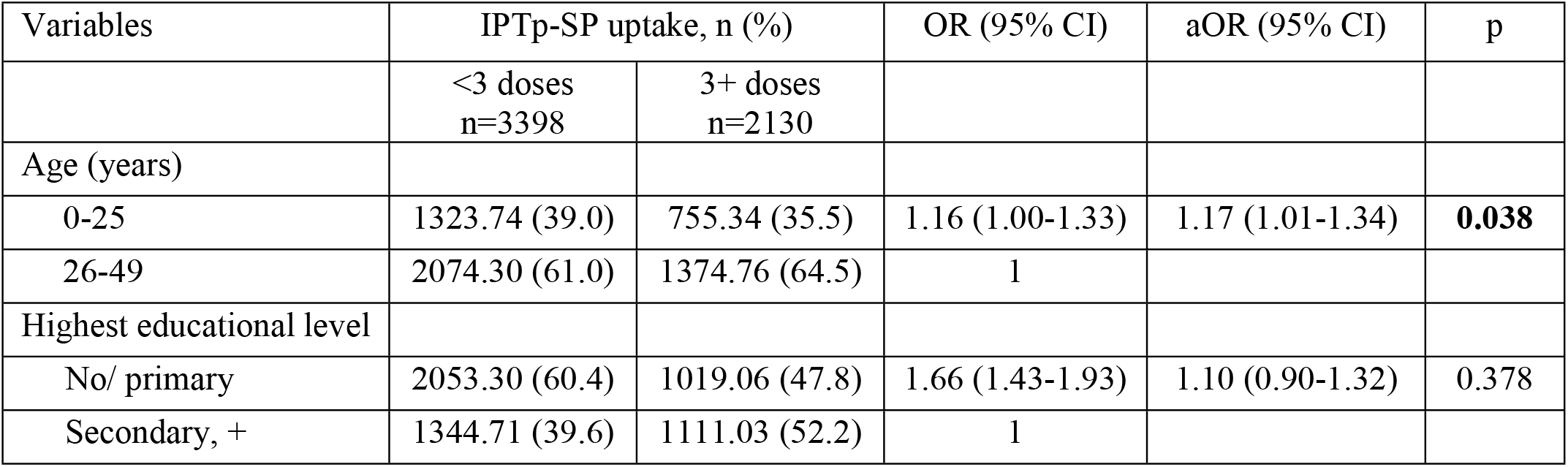
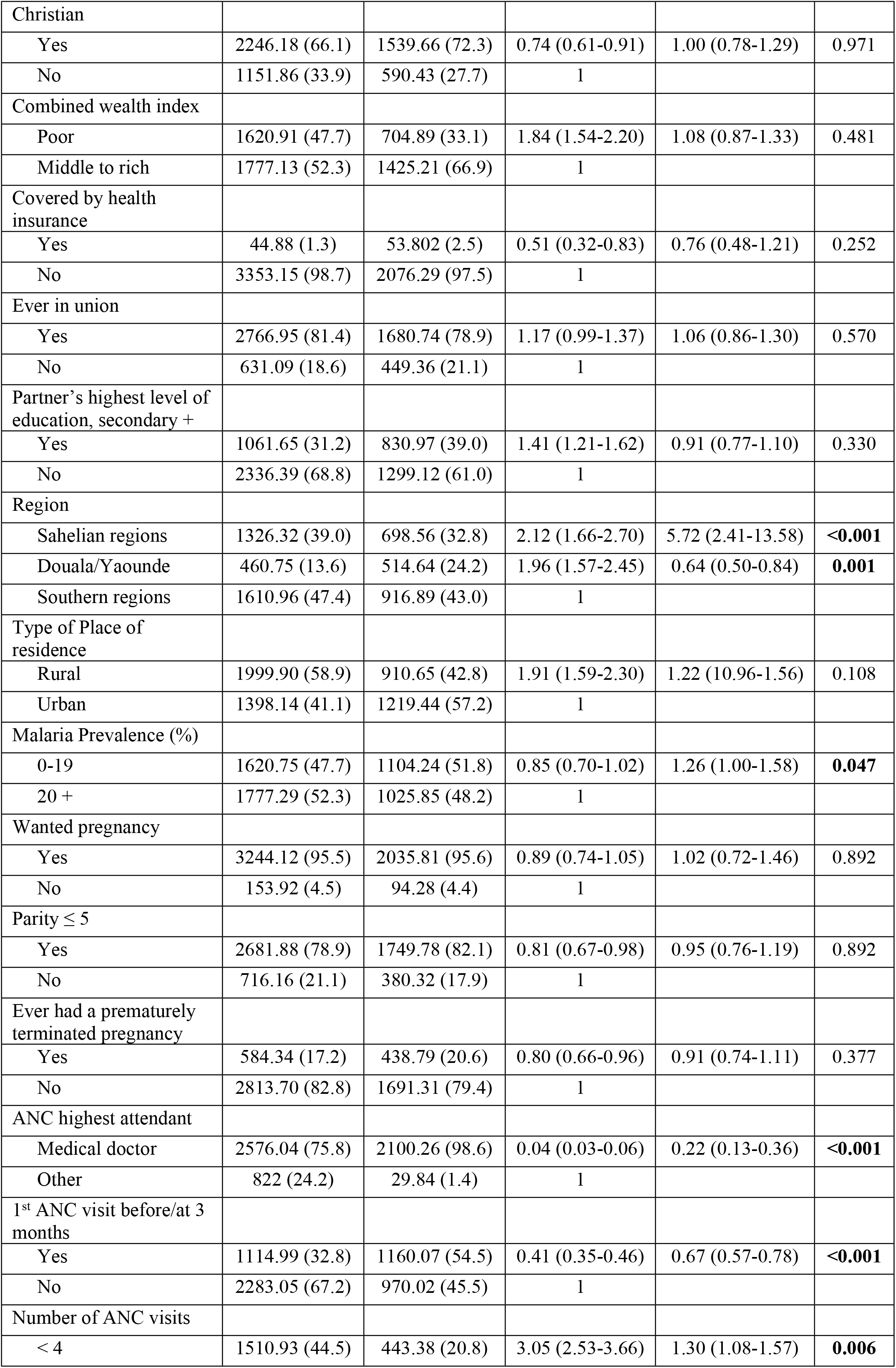
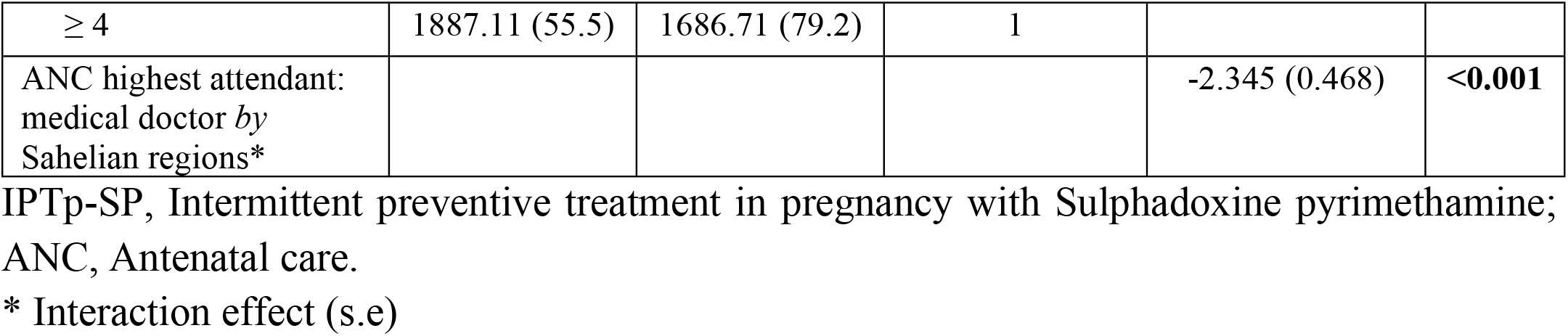

## Discussion

Since 2004 in Cameroon, IPTp-SP stands as a core strategy for the prevention of malaria in pregnancy. We aimed to determine the proportion of, and identify the factors associated with poor uptake of IPTp-SP. We found a high proportion (61.5%) of childbearing age women took less than the recommended ≥3 doses of IPTp-SP during their most recent pregnancy. Factors associated comprised younger maternal age (aOR=1.17, CI: 1.01-1.35), residence in an area of lower endemicity (aOR=1.26, CI: 1.00-1.58), residence in the Sahelian regions (aOR=5.81, CI: 2.46-13.69), and having attended less than 4 ANC visits (aOR=1.30, CI: 1.08-1.57); as well as residence in large cities (aOR=0.65, CI: 0.50-0.84), having attended a first ANC visit at 3 months or less of pregnancy (aOR=0.66, CI: 0.57-0.78) and having been followed on ANC by a medical doctor (aOR=0.22, CI: 0.13-0.35).

In line with the Rollback malaria (RBM) Partnership, Cameroon’s ministry of health targets at 80% the 3 doses coverage of IPTp-SP by 2023 [15]. Though the found 38.5% are a big improvement from the 12.9% in 2011 [10]; the country still has some way to go. However, a 2013 cross-country DHS analytical study found the ≥3 doses coverage of IPTp-SP varying between 0.3% to 43% in sub-Saharan Africa, thus ranking Cameroon among the “high IPTp coverage countries”. Countries with reported lower ≥3 doses coverage of IPTp-SP seem characterized by greater malaria funding, unclear or tardily adopted relevant policy, lower levels of ANC attendance with greater proportion of missed IPTp-SP administration opportunities, and general healthcare system issues (as stock-outs or user fees) [16-19].

In Cameroon as in most sub-Saharan African countries, IPTp-SP distribution was mainly channeled through the ANC program and has become an integral part of it. It ensues from this, as we could find in our study, that women who have their 1st contact with health care providers not early enough and who attend a lower number of subsequent visits are less likely to receive the recommended doses. This concords with studies showing early timing of initial ANC visit to improve uptake of IPTp-SP [17, 20-22]; and those demonstrating an increasing probability of completing IPTp-SP doses with the number of attended ANC visits [10, 11, 23-25]. Nonetheless, only 47.2% of women having attended over 4 visits did take recommended doses. This depicts an important frequency of missed opportunities for IPTp-SP administration. Barriers standing between ANC and uptake of ≥3 doses of IPTp-SP can be broadly grouped into health system-related factors, ANC attendant related factors, maternal related factors. ANC attendant qualification, as found here, can therefore determine the uptake of ≥3 doses of IPTp-SP, with doctors generally providing better care, when compared with other attendants. A study in the northwest reported up to 35.9% of health providers had received no training on IPT and about 30% not knowing when to start the preventive treatment [9], aligning with numerous studies finding a poor general knowledge regarding intermittent preventive treatment among healthcare providers [17].

Researchers equally report numerous maternal-related factors as predictors of poor IPTp3 uptake. We found younger age to be associated with poor uptake of ≥3 doses of IPTp-SP. Age is a rather unsettled factor across countries [17]. Our finding is however consistent with a study in the South-west region of Cameroon, though not statistically significant, supporting that younger women were seemingly showing lower interest for IPTp-SP [12]. A Nigerian study found similar results [21]. Younger women, with less maternal experience, might overlook the subtle symptoms of malaria in pregnancy, or underestimate its effects, thus the lower interest for the preventive treatment.

Our study furthermore shows that women residing in areas of higher endemicity had a higher risk of not taking the three or more recommended doses of IPTp-SP. This supports the result of the DHS multi-country analytical study showing an inverse association between the level of malaria transmission and the uptake of ≥3 doses of IPTp-SP [24]. A potential explanation would be an increase in drug shortage or service overwhelming from the elevated demand or the dodging of preventive doses because of recent treatment received for frequent infections. The efficiency principle however calls for a better consideration of exposure when planning the health initiative.

Also, women residing in the Sahelian regions of the country were found more likely to not taking the recommended ≥3 doses of IPTp-SP compare to those residing in the southern regions; and an opposite association was found for women residing in the great cities (Douala/ Yaounde). The Sahelian regions contains 3 of the 5 regions where women report greater issues with healthcare access [14]. The low level of socio-economic development in the region equally renders healthcare delivery deficient, with the Far North, for example, only containing 8% of Cameroonian doctors despite its 18% of the population [26]. These also give us a glimpse of how the potential gain in women completing their doses of IPTp-SP from an added medical doctor as ANC attendant would be significantly greater when added in the Sahelian regions than in the southern regions; as our cross-level interaction term showed.

## Conclusion

Despite significant improvement with time, coverage of ≥3 doses of IPTp-SP in Cameroon is still low. We identified maternal age, malaria endemicity, place of residence, number of ANC visits attended, timing of first ANC visit, and ANC attendant (which had its effect significantly modified when residing in the Sahelian regions) as important predictors of the uptake of ≥3 doses of IPTp-SP. While pregnant women should be encouraged to strictly follow the ANC policy recommendations, further studies should be carried to evaluate ANC attendant related factors that could explain the elevated proportion of missed opportunities for IPTp-SP administration. Furthermore, the finding of lower IPTp-SP uptake in area of higher endemicity calls for better management of available resources. Greater attention shall be given to the Sahelian regions, where policymakers shall first comprehend the specific obstacles to both healthcare access and service delivery and develop strategies that overcome these obstacles.

## Data Availability

https://doi.org/10.6084/m9.figshare.21308949

https://figshare.com/articles/dataset/Finaldatabase_DHSIPTpSPAnalysis_sav/21308949

## Acknowledgments

We gratefully thank *The DHS Program* team, who has granting us access to the 2018 Cameroon Demographic Health Survey database.

